# SARS-CoV-2 Spike S1-specific IgG kinetic profiles following mRNA- versus vector-based vaccination in the general Dutch population

**DOI:** 10.1101/2021.10.25.21265467

**Authors:** Lotus L. van den Hoogen, Marije K. Verheul, Eric R.A. Vos, Cheyenne C.E. van Hagen, Michiel van Boven, Denise Wong, Alienke J. Wijmenga-Monsuur, Gaby Smit, Marjan Kuijer, Debbie van Rooijen, Marjan Bogaard-van Maurik, Ilse Zutt, Jeffrey van Vliet, Fiona R.M. van der Klis, Hester E. de Melker, Robert S. van Binnendijk, Gerco den Hartog

## Abstract

mRNA- and vector-based vaccines are used at a large scale to prevent COVID-19. We compared Spike S1-specific (S1) IgG antibodies after vaccination with mRNA-based (Comirnaty, Spikevax) or vector-based (Janssen, Vaxzevria) vaccines, using samples from a Dutch nationwide cohort. mRNA vaccines induced faster inclines and higher S1 antibodies compared to vector-based vaccines in adults 18-64 years old (n=2,412). For all vaccines, one dose resulted in boosting of S1 antibodies in adults with a history of SARS-CoV-2 infection. For Comirnaty, two to four months following the second dose (n=196), S1 antibodies in adults aged 18-64 years old (436 BAU/mL, interquartile range: 328-891) were less variable and median concentrations higher compared to those in persons ≥80 years old (366, 177-743), but differences were not statistically significant (p>0.100). Nearly all participants seroconverted following COVID-19 vaccination, including the aging population. These data confirm results from controlled vaccine trials in a general population, including vulnerable groups.

## Introduction

High vaccine effectiveness (VE) data related to the prevention of hospitalization due to COVID-19 have been reported up to 20 weeks after vaccination.^1,2^ In the Netherlands, four vaccines have been included in the national vaccination program and are either mRNA-based (Comirnaty and Spikevax) or vector-based (Vaxzevria and Janssen). mRNA-based vaccines use lipid nanoparticles to deliver Spike-encoding mRNA and vector-based vaccines use adenovirus to deliver Spike-encoding DNA to induce expression of the Spike protein by human cells. Multiple reports independently describe the induction of Spike-specific antibodies by the various vaccines. However, most results are from specific groups such as immunocompromised patients or healthcare workers.^3-5^ Direct comparisons of mRNA- and vector-based vaccines in the general population are scarce as well as those comparing these four vaccines simultaneously.^6^

PIENTER-Corona (PiCo) is an ongoing cohort study in a Dutch nationwide sample at four-monthly intervals.^7,8^ Using this cohort, we selected participants 18 years or older who had received one or two doses of Comirnaty, Spikevax, Vaxzevria or Janssen and compared SARS-CoV-2 Spike S1-specific (S1) antibody concentrations induced by each vaccine type.

## Results

The two most recent PiCo study rounds in February and June 2021 followed the launch of the Dutch national COVID-19 vaccination campaign. Data availability was driven by vaccine roll-out and prioritization of the aging population during the campaign (Sup Fig 1). For those 18-64 years old, data were available up to two months following each dose across all four vaccines (n=2,412), while for Comirnaty, data across all ages were available in SARS-CoV-2-naïve persons up to four months following the second dose (18-91 years old, n=196). S1 antibody concentrations were measured using a previously described assay and expressed in binding antibody units per mL (BAU/mL), as defined by the WHO International Standard (NIBSC 20/136).^9,10^ SARS-CoV-2 infection prior to vaccination was defined by S1 seropositivity in any study round prior to vaccination or reporting a PCR/antigen SARS-CoV-2 positive test prior to vaccination.

Among adults aged 18-64, prioritization resulted in the fact that participants were relatively frequently healthcare worker or had comorbidities, and most persons were in the oldest age group (45-64 years old) (Table 1). The median vaccination interval between the two doses was 77 days for Vaxzevria (IQR: 69-77) and 35 days (28-35) and 33 days (28-35) for Comirnaty and Spikevax, respectively.

**Table 1:** General characteristics of SARS-CoV-2 vaccinated adults aged 18-64 years up to two months following the first or second dose by vaccination brand and number of doses (n=2,412).

| n (%), unless<br>otherwise specified | **Comirnaty** |  | **Spikevax** |  | **Vaxzevria** |  | **Janssen** |
| --- | --- | --- | --- | --- | --- | --- | --- |
| **Doses** | **1** | **2** | **1** | **2** | **1** | **2** | **1** |
| N | 974 | 466 | 222 | 124 | 111 | 298 | 217 |
| Sex |  |  |  |  |  |  |  |
| - Male | 407 (42%) | 154 (33%) | 87 (39%) | 31 (25%) | 47 (42%) | 98 (33%) | 82 (38%) |
| - Female | 567 (58%) | 312 (67%) | 135 (61%) | 93 (75%) | 64 (58%) | 200 (67%) | 135 (62%) |
| Age in years |  |  |  |  |  |  |  |
| - 18-29 | 68 (7%) | 47 (10%) | 18 (8%) | 11 (9%) | 3 (3%) | 15 (5%) | 26 (12%) |
| - 30-44 | 332 (34%) | 101 (22%) | 78 (35%) | 49 (40%) | 5 (5%) | 40 (13%) | 49 (23%) |
| - 45-64 | 574 (59%) | 318 (68%) | 126 (57%) | 64 (52%) | 103 (93%) | 243 (82%) | 142 (65%) |
| Healthcare worker |  |  |  |  |  |  |  |
| - No | 841 (86%) | 297 (64%) | 184 (83%) | 57 (46%) | 97 (87%) | 179 (60%) | 157 (72%) |
| - Yes | 133 (14%) | 169 (36%) | 38 (17%) | 67 (54%) | 14 (13%) | 119 (40%) | 60 (28%) |
| Comorbidities |  |  |  |  |  |  |  |
| - Risk group* | 236 (24%) | 193 (41%) | 50 (23%) | 39 (31%) | 48 (43%) | 106 (36%) | 32 (15%) |
| - None/other** | 738 (76%) | 273 (59%) | 172 (77%) | 85 (69%) | 63 (57%) | 192 (64%) | 185 (85%) |
| Infection history |  |  |  |  |  |  |  |
| - No | 791 (81%) | 408 (88%) | 175 (79%) | 108 (87%) | 101 (91%) | 265 (89%) | 186 (86%) |
| - Yes | 183 (19%) | 58 (12%) | 47 (21%) | 16 (13%) | 10 (9%) | 33 (11%) | 31 (14%) |
| Median vaccination interval in days (IQR) |  | 35 (28 – 35) |  | 33 (28 - 35) |  | 77 (69 – 77) |  |

\* High risk comorbidities: asthma or other lung disease, cardiovascular disease, diabetes, (history of) cancer, history of transplantation, kidney disease, immune disease, splenectomy, liver disease, rheumatoid arthritis, intestinal disease, neurological disease, or other (open field). Persons with these comorbidities were prioritized during the vaccination campaign in the Netherlands as they were considered high risk for severe COVID-19.
\*\*Other comorbidities: hay fever, skin disease or allergies.
IQR: interquartile range.

In infection-naïve adults aged 18-64 years, mRNA-based vaccines induced S1 IgG faster and reached higher levels than vector-based vaccines (Figure 1). Subsequent to the rise following the second dose, an initial rapid decay could be seen which stabilized, while for vector-based vaccines the slower rise stabilized without any clear decay. Between fourteen days and two months after completion of the vaccination schedule or positive SARS-CoV-2 test, median IgG levels were 2,799 BAU/mL for Spikevax (IQR: 1,714-4,669; seropositivity: 99%, n/N: 72/73), 2,408 for Comirnaty (1,373 -3,799; 99%, 151/152), 313 for Vaxzevria (145-703; 100%, 185/186), 64 for Janssen (29-143; 95%, 189/199), and 91 (39-230; 87%, 90/104) for unvaccinated, SARS-CoV-2-confirmed participants. All three non-responders for Spikevax, Comirnaty and Vaxzevria had a high risk comorbidity (see footnote Table 1). For Janssen, twenty-eight days after vaccination S1 IgG increased to 77 (37-163; 98%, 85/87) with no comorbidity for the two non-responders. Regression results showed that age, sex, and comorbidity significantly contributed to S1 IgG concentrations but this was not consistent between vaccines and doses (Sup Table 1).

**Figure 1:**
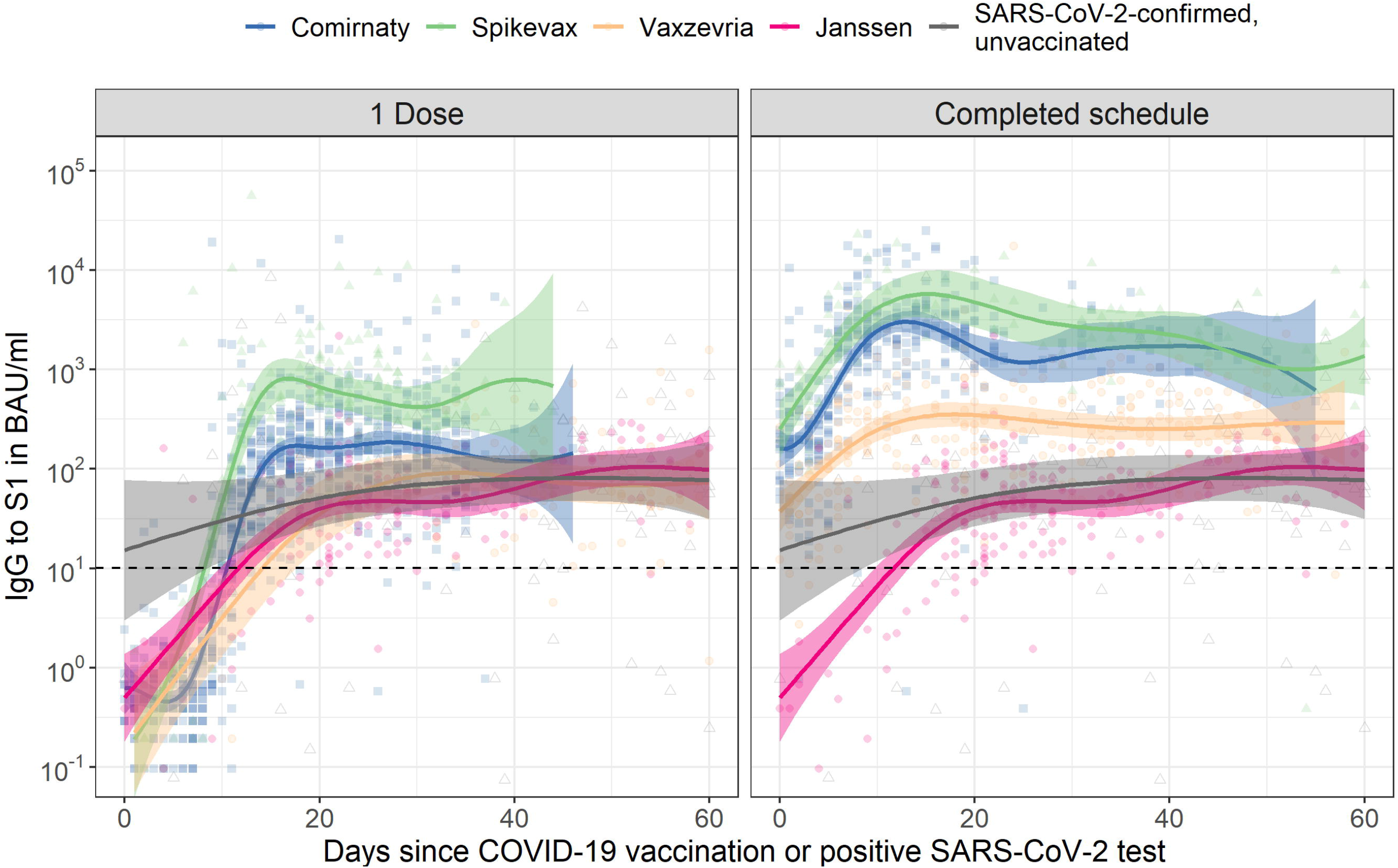
Spike S1 immunoglobulin G (IgG) kinetics following COVID-19 vaccination by number of doses and vaccine brand in SARS-CoV-2-naive adults aged 18 to 64 years old. The dashed horizontal line represents the threshold for seropositivity. Data for Janssen is duplicated across the two panels to enable direct comparison with the other vaccine brands after one dose and a completed vaccination schedule. For comparison, IgG concentrations following a positive SARS-CoV-2 PCR or antigen test in unvaccinated participants are shown alongside vaccination responses; data is duplicated in both panels (for details see Supplementary Table 3). Fit and 95% confidence bands are shown from a Generalized Additive Model, using penalized splines, with only time since dose in days as explanatory variable. For results of multivariable models, see Supplementary Table 1. BAU/mL: binding antibody units per mL; IgG: immunoglobulin G.

In adults aged 18-64, S1 IgG concentrations were higher in persons with a history of SARS-CoV-2 after one vaccine dose compared to previously naive persons after a completed schedule irrespective of vaccine type, Figure 2A. For persons with an infection history, no further increases were seen after a second dose if applicable (p>0.100).

**Figure 2:**
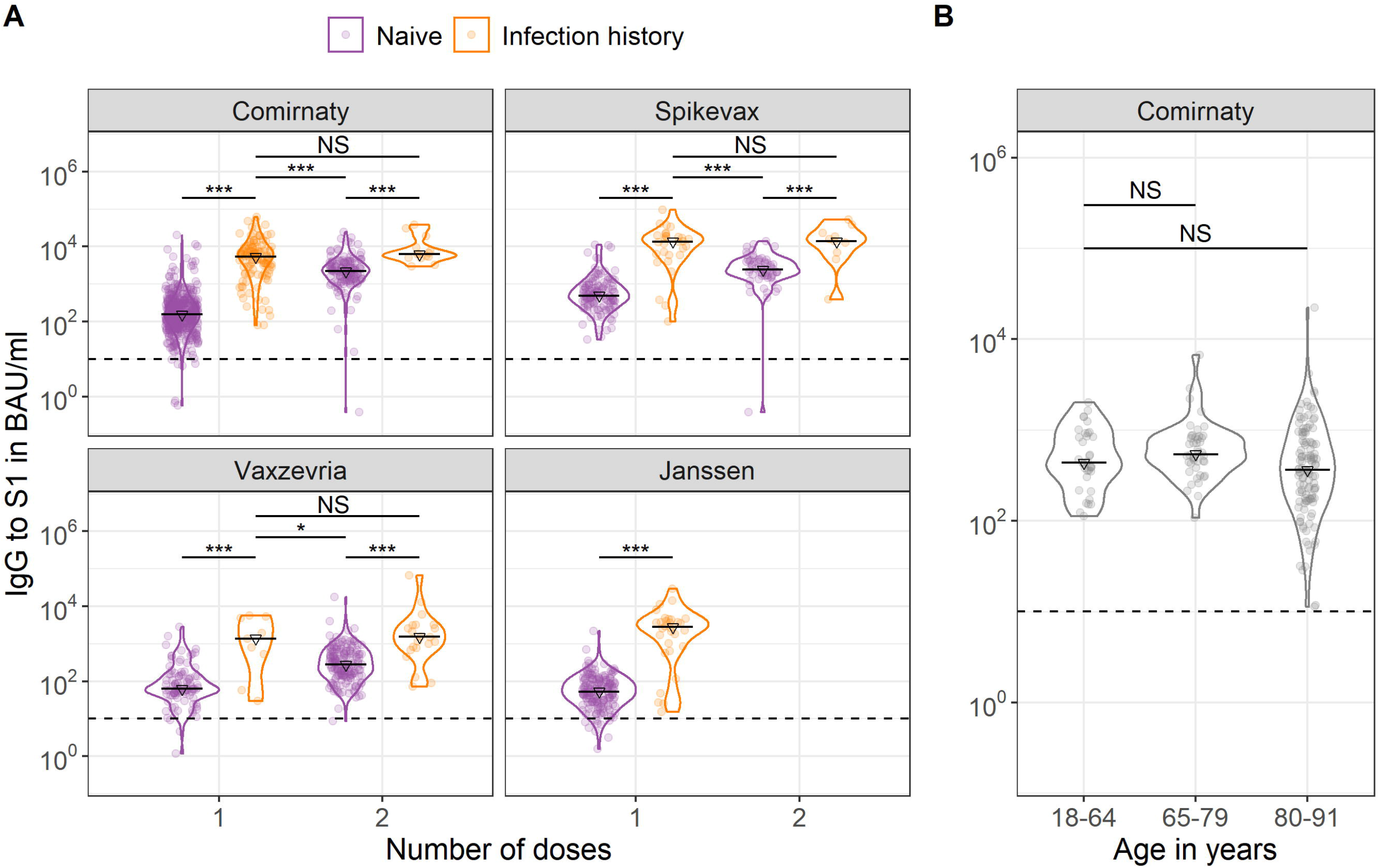
Violin plots of Spike S1 immunoglobulin G (IgG) concentrations by number of COVID-19 vaccine doses, SARS-CoV-2 infection history and vaccine brand (A) and in SARS-CoV-2-naïve adults at two to four months following a completed Comirnaty schedule by age group (B). Triangles and black horizontal lines represent median concentrations of IgG to Spike S1 in BAU/mL. The dashed horizontal line represents the threshold for seropositivity. In (A) IgG measurements were taken between two weeks and two months after the indicated dose; while in (B) between two and four months after completion of the Comirnaty schedule. Wilcoxon-Mann-Whitney tests were used to test for differences in IgG concentrations by infection history in (A) with blocking for strata of sex, and by age group in (B) with blocking for strata of sex and time since second dose (less versus more than three months). ^*^ p<0.05; ^**^ p<0.01; ^***^ p <0.001; NS: not significant (p>0.100); BAU/mL: binding antibody units per mL; IgG: immunoglobulin G.

For Comirnaty, IgG levels were more heterogeneous in the oldest age group but did not show a statistically significant difference between infection-naïve adults aged 18-64 years (436, 328-891) and those aged 65-79 (542, 417-836) and ≥80 years old (366, 177-743), p>0.100, (Figure 2B, Sup Table 2).

## Discussion

In response to the global COVID-19 pandemic, novel vaccine strategies using mRNA- and vector-based induction of the immune system have been developed. In adults aged 18 to 64, we observed the steepest inclines and highest Spike S1 IgG concentrations up to two months following vaccination with mRNA-based vaccines compared to vector-based COVID-19 vaccines. Boosting of Spike S1 IgG responses in persons with a history of infection was seen for all four vaccines, and a second dose did not further increase anti-S1 IgG levels. We used antibody concentration units according to the WHO international standard, enabling direct comparison between studies.

Others have shown that IgG to Spike S1 following SARS-CoV-2 infection or mRNA vaccination shows an initial decline with stabilization of IgG antibodies after 4-6 months.^11^ The rapid induction of high levels of antibodies by mRNA vaccines, compared to vector-based vaccines, followed by an early decay may point to the induction of short-lived plasma blasts by the mRNA vaccines that disappear soon after the immunization, and may not predict the number of sustaining memory cells.

During the delta variant period in the Netherlands, overall VE against hospitalization was 91% for Comirnaty, 96% for Spikevax, 88% for Vaxzevria and 82% for Janssen.^1^ Antibody data from our study mirrored these trends which supports the notion that antibody binding and neutralization correlate with vaccine efficacy.^12^ However, the relative difference in VE estimates is smaller than in antibody levels, indicating that antibody levels alone do not constitute immune protection, as expected.

Boosting of S1 IgG after one dose of mRNA or vector vaccine in previously infected persons has been described previously.^5,13^ Here we confirm boosting of infection-induced immune memory regardless of vaccine-type. This boosting seems stronger than observed after revaccination, indicating a more matured underlying memory B cell response induced by viral infection compared to immunization. This observation needs in-dept follow-up in the future when the opposite will occur: boosting of vaccine-induced immunity by infection.

We showed highly variable antibody response between individuals which increased in community-dwelling elderly aged 80 years and older after completion of Comirnaty. To date, most of the humoral data in the elderly are from nursing home residents.^14^ Müller et al. showed lower antibody profiles in nursing home residents compared to those 20-60 year old shortly following vaccination, which contrast with our findings. Such discrepancies might be caused by the increased age range (up to 91 in our study versus 100 in Müller et al.), presence of more complex comorbidities or the fact that antibody production is delayed in the elderly.

The data presented here are a highly relevant confirmation of results from controlled vaccine trials since we show high immunogenicity after vaccination in the general population, including vulnerable groups and different vaccination regimens. Although most persons seroconverted regardless of the vaccine received, mRNA- and vector-based COVID-19 vaccines induced distinct S1-specific IgG kinetic profiles. Further exploration of the translation of antibody quantity to antibody quality and subsequent protection against infection and (severe) disease as well as the involvement of other immune compartments such as T cells is needed.

## Methods

### Study population

We used samples from a four-monthly prospective nationwide cohort study in the Netherlands which has been described in detail elsewhere.^7,8^ Participants provided a fingerprick blood sample and completed a questionnaire including sociodemographic factors, comorbidities, COVID-19 disease (symptoms, type and date of SARS-CoV-2 test) and COVID-19 vaccination (brand and dates). Participants 18 years or older who had received one or two doses of Comirnaty, Spikevax, Vaxzevria or Janssen were selected. Unvaccinated participants aged 18-64 years old who reported a positive SARS-CoV-2 PCR or antigen test up to two months prior to sampling were selected to compare vaccine responses to those following SARS-CoV-2 infection (n=114; Sup Table 3).

### Antibody detection

Serum samples were analyzed for IgG concentrations to SARS-CoV-2 Spike S1 using a previously described bead-based assay.^9^ IgG concentrations were expressed in binding antibody units (BAU/mL) using the NIBSC 20/136 WHO standard.^10^

### Statistical analyses

Statistical analyses were done in R Studio (version 4.1.0).^15^ Mann-Whitney tests were used to compare IgG concentrations by age group with blocking per strata of sex and time since vaccination (dichotomized as more or less than 3 months), and by infection history with blocking per strata of sex (*coin*^16^). IgG kinetics were fitted with a Generalized Additive Model, using penalized splines (*mgcv*^17^).

## Data Availability

All data produced in the present study are available upon reasonable request to the authors

## Acknowledgements

We would like to thank all participants.

