## Supplementary Figure and Tables for "SARS-CoV-2 Spike S1-specific IgG kinetic profiles following mRNA- versus vector-based vaccination in the general Dutch population"

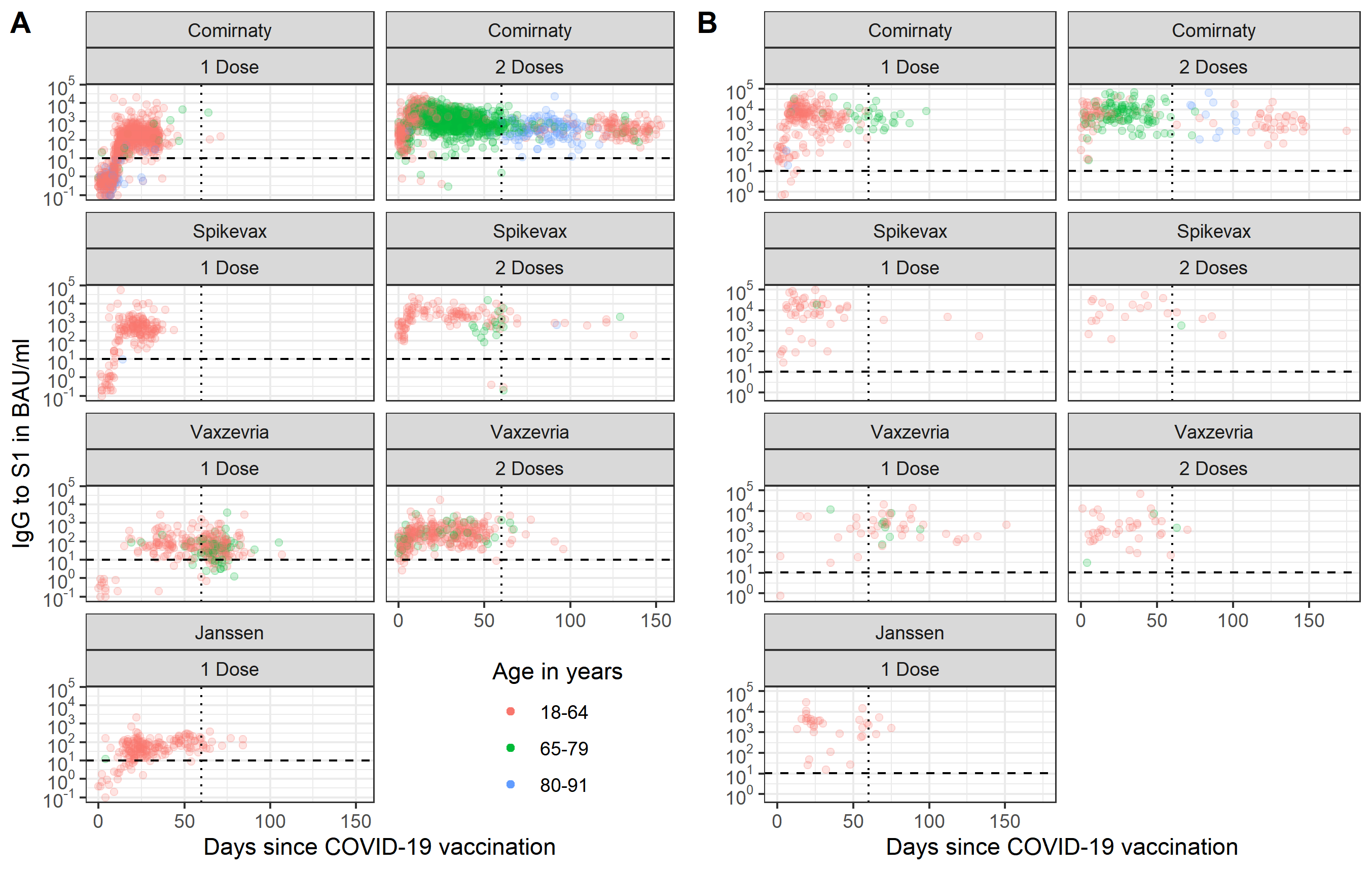
**Supplementary Material**

**Supplementary Figure 1: Spike S1 immunoglobulin G (IgG) concentrations over time by age group, number of doses and vaccine brand for SARS-CoV-2-naive participants (A) and participants with a SARS-CoV-2 infection history (B).** Measurements for a total of 4,110 participants are shown; data left of the dotted vertical line at two months was included in the analyses for adults aged 18-64 years old to enable direct comparison of vaccines (n=2,412). For Comirnaty, data was also analyzed between two and four months following the second dose in SARS-CoV-2-naïve participants across all ages (18-91 years old; n=196). The dashed horizontal line represents the threshold for seropositivity. BAU/mL: binding antibody units per mL; IgG: immunoglobulin G.

**Supplementary Table 1: Generalized Additive Model regression results for Spike S1 immunoglobulin G (IgG) concentrations per vaccine type and dose in the SARS-CoV-2-naïve study population aged 18-64 years up to two months following the indicated dose.** IgG concentrations in BAU/ml were log10-transformed. All covariates were included in the initial model (time since dose in days, age group, sex and comorbidity) and the final model was created using backward selection by manually dropping the variable with the highest p-value at each step and selecting the more complex model if it resulted in a decrease in Akaike’s Information Criterion (AIC) of at least two. No interactions were considered.

|  | **Comirnaty** | | **Spikevax** | | **Vaxzevria** | | **Janssen** | | **Comirnaty** | | | **Spikevax** | | **Vaxzevria** | |
| --- | --- | --- | --- | --- | --- | --- | --- | --- | --- | --- | --- | --- | --- | --- | --- |
| **Doses** | **1 Dose** | | | | | | | | | **2 Doses** | | | | | |
| **N** | 791 | | 175 | | 101 | | 186 | | 408 | | | 108 | | 265 | |
|  | Coef. | p-value | Coef. | p-value | Coef. | p-value | Coef. | p-value | Coef. | | p-value | Coef. | p-value | Coef. | p-value |
| **Age in years** |  |  |  |  |  |  |  |  |  | |  |  |  |  |  |
| 18-29 | Ref |  |  |  |  |  |  |  | Ref | |  |  |  |  |  |
| 30-44 | -0.30 | <0.001 |  |  |  |  |  |  | -0.28 | | 0.007 |  |  |  |  |
| 45-64 | -0.42 | <0.001 |  |  |  |  |  |  | -0.30 | | 0.002 |  |  |  |  |
| **Sex** |  |  |  |  |  |  |  |  |  | |  |  |  |  |  |
| Male | Ref |  | Ref |  |  |  | Ref |  | Ref | |  |  |  |  |  |
| Female | 0.15 | <0.001 | 0.23 | 0.023 |  |  | 0.21 | 0.006 | 0.19 | | 0.001 |  |  |  |  |
| **Comorbidity** |  |  |  |  |  |  |  |  |  | |  |  |  |  |  |
| None/other* |  |  | Ref |  | Ref |  |  |  |  | |  |  |  | Ref |  |
| High risk** |  |  | -0.30 | 0.019 | -0.22 | 0.045 |  |  |  | |  |  |  | -0.17 | 0.005 |
| **Time since dose in days (spline)** |  | <0.001 |  | <0.001 |  |  |  | <0.001 |  | | <0.001 |  | <0.001 |  | <0.001 |

*Other comorbidities: hay fever, skin disease or allergies.
******High risk comorbidities: asthma or other lung disease, cardiovascular disease, diabetes, (history of) cancer, history of transplantation, kidney disease, immune disease, splenectomy, liver disease, rheumatoid arthritis, intestinal disease, neurological disease, or other (open field). Persons with these comorbidities were prioritized during the vaccination campaign in the Netherlands as they were considered high risk for severe COVID-19.
BAU/mL: binding antibody units per mL; IgG: immunoglobulin G; Coef: coefficient; Ref: reference.

**Supplementary Table 2: General characteristics of the SARS-CoV-2-naïve study population between two and four months after completion of the Comirnaty vaccination schedule by age category.**

|  | **18 to 64** | **65 to 79** | **80 to 91** |
| --- | --- | --- | --- |
| N | 35 | 44 | 117 |
| Sex |  |  |  |
| - Male | 11 (31%) | 27 (61%) | 70 (60%) |
| - Female | 24 (69%) | 17 (39%) | 47 (40%) |
| Healthcare worker |  |  |  |
| - No | 8 (23%) | 42 (95%) | 117 (100%) |
| - Yes | 27 (77%) | 2 (5%) | 0 (0%) |
| Comorbidities |  |  |  |
| - Risk group* | 15 (43%) | 24 (55%) | 63 (54%) |
| - None/other** | 20 (57%) | 20 (45%) | 54 (46%) |
| Median vaccination interval in days (IQR) | 28 (25 – 35) | 35 (35 – 36) | 35 (35 – 37) |
| Median time since second dose in days (IQR) | 104 (83 – 116) | 69 (63 – 80) | 86 (75 – 96) |

*****High risk comorbidities: asthma or other lung disease, cardiovascular disease, diabetes, (history of) cancer, history of transplantation, kidney disease, immune disease, splenectomy, liver disease, rheumatoid arthritis, intestinal disease, neurological disease, or other (open field). Persons with these comorbidities were prioritized during the vaccination campaign in the Netherlands as they were considered high risk for severe COVID-19.
**Other comorbidities: hay fever, skin disease or allergies.
IQR: interquartile range.

**Supplementary Table 3: General characteristics of the unvaccinated adults aged 18-64 years up to two months following a positive SARS-CoV-2 test.** Participants who reported a positive PCR or antigen test for SARS-CoV-2 up to two months prior to sampling were included.

|  | **n (%)** |
| --- | --- |
| N | 114 |
| Sex |  |
| - Male | 34 (30%) |
| - Female | 80 (70%) |
| Age in years |  |
| - 18-29 | 27 (24%) |
| - 30-44 | 37 (32%) |
| - 45-64 | 50 (44%) |
| Healthcare worker |  |
| - No | 77 (68%) |
| - Yes | 37 (32%) |
| Comorbidities |  |
| - Risk group* | 28 (25%) |
| - None/other** | 86 (75%) |
| COVID-19 symptoms when tested |  |
| - No | 14 (12%) |
| - Yes | 100 (88%) |
| Median time since positive SARS-CoV-2 test in days (IQR) | 40 (24 – 52) |

***** High risk comorbidities: asthma or other lung disease, cardiovascular disease, diabetes, (history of) cancer, history of transplantation, kidney disease, immune disease, splenectomy, liver disease, rheumatoid arthritis, intestinal disease, neurological disease, or other (open field). Persons with these comorbidities were prioritized during the vaccination campaign in the Netherlands as they were considered high risk for severe COVID-19.
**Other comorbidities: hay fever, skin disease or allergies.
IQR: interquartile range, PCR: polymerase chain reaction.
